# Public Opinion about the UK Government during COVID-19 and Implications for Public Health: A Topic Modelling Analysis of Open-Ended Survey Response Data

**DOI:** 10.1101/2021.03.24.21254094

**Authors:** Liam Wright, Alexandra Burton, Alison McKinlay, Andrew Steptoe, Daisy Fancourt

## Abstract

Confidence in the central UK Government has declined since the beginning of the COVID-19 pandemic, and while this may be linked to specific government actions to curb the spread of the virus, understanding is still incomplete. Examining public opinion is important, as research suggests that low confidence in government increases the extent of non-compliance with infection-dampening rules (for instance, social distancing); however, the detailed reasons for this association are still unclear. To understand public opinion on the central UK government during COVID-19, we used structural topic modelling, a text mining technique, to extract themes from over 4000 free-text survey responses, collected between 14 October and 26 November 2020. We identified eleven topics, among which were topics related to perceived government corruption and cronyism, complaints about inconsistency in rules and messaging, lack of clear planning, and lack of openness and transparency. Participants reported that elements of the government’s approach had made it difficult to comply with guidelines (e.g., changing rules) or were having impacts on mental wellbeing (e.g., inability to plan for the future). Results suggested that consistent, transparent communication and messaging from the government is critical to improving compliance with measures to contain the virus, as well as protecting mental health during health emergencies.

## Introduction

The pandemic of COVID-19 has disrupted lives across the globe. Besides the impact on public health, the economic and social costs have been substantial. Governments have been tasked with balancing limiting infection rates against the competing goals of maintaining civic freedoms and supporting jobs and economic production. Policy decisions have been made against a backdrop of significant uncertainty in a rapidly evolving, often unclear situation (Smith et al., 2020). Governments have differed markedly in the strategies that they have adopted (Hale et al., 2020), and public opinion about governments’ handling of the pandemic has varied widely (YouGov, 2021). Opinion has differed not only on the effectiveness of measures implemented, but also on specific actions, such as the extent to which governments have supported vulnerable groups (Lazarus et al., 2020).

As of February 2021, the UK has been one of the countries hardest hit by COVID-19, as measured by infection rates, deaths and lost economic production (John Hopkins University, 2021; OECD, 2020). Confidence in the UK central government’s handling of the pandemic has overall been low (Duffy, 2020; YouGov, 2021). While confidence has increased in recent months (YouGov, 2021), it has declined from the start of the pandemic and to a greater extent than for the devolved governments of Wales and Scotland (Fancourt, Bu, et al., 2020; also see Supplementary Figure S1). Some of the decrease in confidence has been linked to specific policy actions, such as the decision to keep a senior government advisor, Dominic Cummings, in post after reports he broke lockdown rules (Fancourt, Steptoe, et al., 2020). But gradual changes in confidence have occurred since then (Fancourt, Bu, et al., 2020; YouGov, 2021). Whilst these trends have been well mapped, little is known on the specifics of public opinion such as which aspects of the government’s response have caused most concern amongst the public. Additionally, it is widely acknowledged that maintaining public confidence is imperative during pandemics, as trust and confidence are related to increased compliance with social distancing and other infection-dampening rules (Brouard et al., 2020; Coroiu et al., 2020; Wright et al., 2020), but it remains unclear how and why public opinions might be related to preventive behaviours. It is also how government actions have been related to individuals’ health and wellbeing during COVID-19, more generally.

This research gap is complex given methodological challenges in assessing public opinions. Closed-ended questions constrict responses to categories that researchers have determined in advance (Geer, 1991). Although open-ended questions can increase non-response and bias the sample towards more articulate respondents (Geer, 1988), obtaining spontaneous responses is arguably particularly important in the current pandemic as there is little prior data or experience to predict how the public is reacting to the government’s performance. While analysing open-ended survey data has historically posed problems for research, requiring resource-intensive manual coding and typically involving small sample sizes, the development of text mining techniques and accompanying software tools has enabled the timely quantitative analysis of large-scale open-ended survey data. Therefore, in this paper, we used structural topic modelling (STM; Roberts et al., 2014) to analyse open-ended survey data from 4,000 UK adults during the COVID-19 pandemic. We used STM to explore opinions on the UK government’s responses to the pandemic, test whether opinions differ according to participant characteristics, investigate how these opinions cluster together, and consider how they relate to public health.

## Methods

### Participants

We used data from the COVID-19 Social Study; a large panel study of the psychological and social experiences of over 70,000 adults (aged 18+) in the UK during the COVID-19 pandemic. The study commenced on 21 March 2020 and involved online weekly data collection for 22 weeks with monthly data collection thereafter. The study is not random and therefore is not representative of the UK population, but it does contain a heterogeneous sample. Full details on sampling, recruitment, data collection, data cleaning and sample demographics are available at https://github.com/UCL-BSH/CSSUserGuide. The study was approved by the UCL Research Ethics Committee [12467/005] and all participants gave informed consent.

A one-off free-text module was included in the survey between 14 October and 26 November. Participants were asked to write responses to a range of questions about their experiences during the pandemic and their expectations for the future. These questions were purposefully general in order to be useful for several research studies. We used responses from five questions (Table 1) that were considered to have the potential to elicit opinions about the central UK Government. Given that participants were not explicitly asked to discuss the government, we only analysed responses that included keywords related to the government (e.g., minister, government, politician). Keywords were identified by searching through the list of unique words from all responses and are displayed in the Supplementary Information. We only analysed responses from residents of England to focus on the central UK government. A flow chart of the sample selection is displayed in Figure 1. A description of the political context over the study period is provided in the Supplementary Information.

**Table 1:**
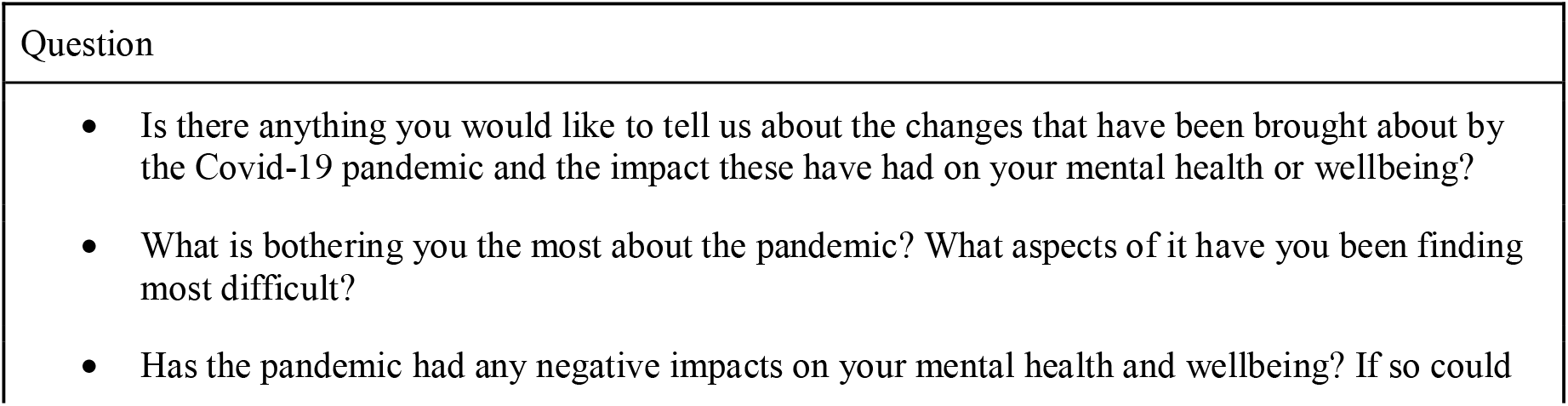
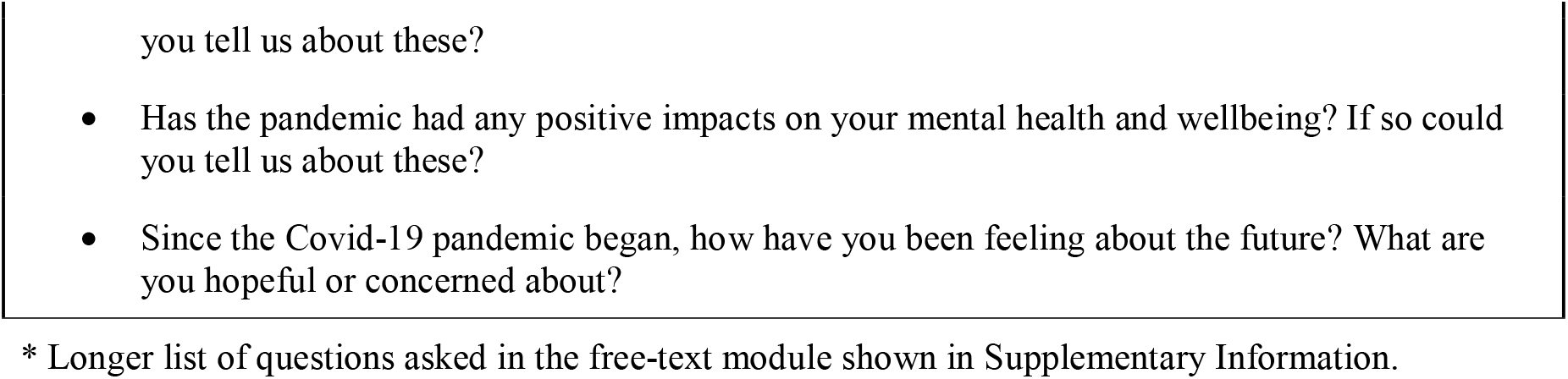
Free-Text Response Questions.

**Figure 1.**
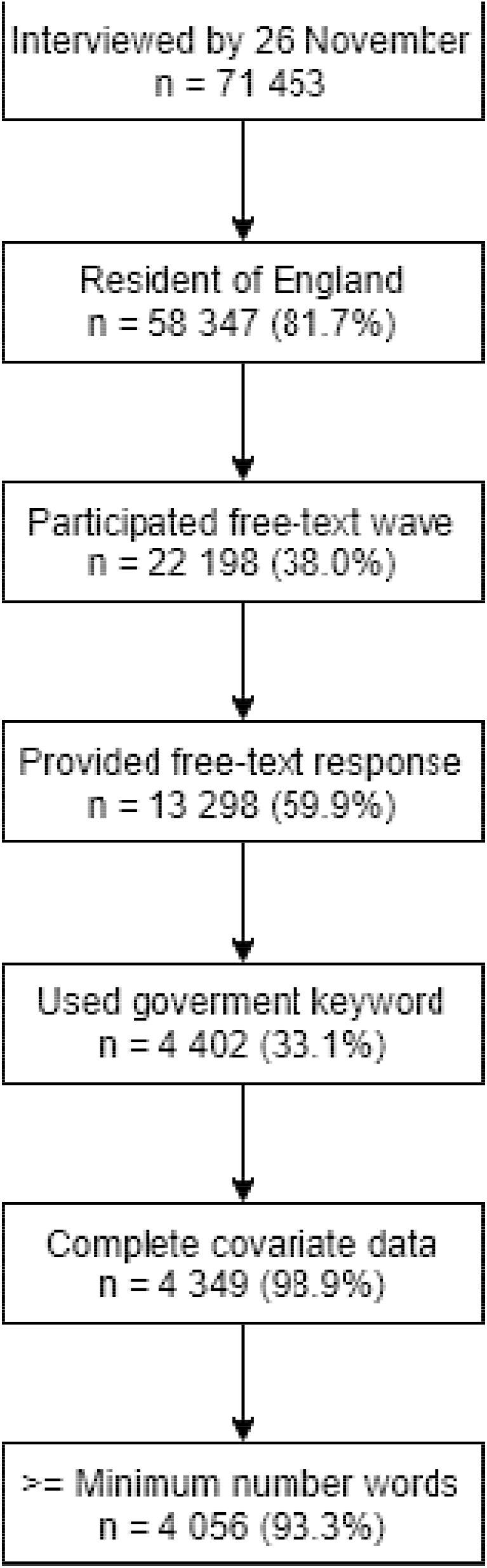
Flow diagram

### Data cleaning

We performed topic modelling using unigrams (single words). Responses were cleaned using an iterative process, described in detail in the Supplementary Information. To reduce data sparsity, we used word stemming using the Porter (1980) algorithm. Data cleaning was carried out in R version 3.6.3 (R Core Team, 2020). The code used is available at https://osf.io/jw3gb/. The data are not available due to stipulations set out by the ethics committee.

### Data analysis

We performed several quantitative analyses. First, we ran a Heckman selection probit model to explore the predictors of whether a respondent provided a free-text response and whether, conditional on having provided a response, the participant used a government keyword. Second, we used STM, implemented with the stm R package (Roberts et al., 2019), to extract topics from responses that used government keywords. We treated as a single document all answers given by a respondent that included one or more government keywords. STM treats documents as a probabilistic mixture of topics and topics as a probabilistic mixture of words. It is a “bag of words” approach that uses correlations between word frequencies within documents to define topics. STM allows for inclusion of covariates in the estimation model, such that the estimated proportion of a text devoted to a topic can differ according to document metadata (e.g., characteristics of its author). We included participant’s gender, ethnicity (white, non-white), age (grouped: 18-29, 30-44, 45-59, 60+), education level (degree or above, A-Level or equivalent, GCSE or below), employment status (employed, inactive or retired, student, unemployed), each collected at first data collection, and date of response, confidence in government (How much confidence do you have in the UK GOVERNMENT that they can handle Covid-19 well? 1 “None at all” – 7 “Lots”) and adherence to COVID-19 guidelines (Are you following the recommendations from authorities to prevent spread of Covid-19? 1 “Not at all” −7 “Very much so”), collected during the same data collection as the free-text responses. There was only a very small amount of item missingness, so we used complete case data.

We ran STM models from 2-60 topics and selected the final model based on visual inspection of the semantic coherence and exclusivity of the topics and close reading of exemplar documents representative of each topic. Semantic coherence measures the degree to which high probability words within a topic co-occur, while exclusivity measures the extent to that a topic’s high probability words have low probability for other topics. We erred on the side of choosing fewer topics, given the sample size in our study.

After selecting a final model, we carried out three further analyses. First, we decided upon narrative descriptions for the topics based on high probability words, high “FREX” words (a weighted measure of word frequency and exclusivity), and exemplar texts (responses with a higher proportion of text on a given topic). Second, we ran regression models estimating whether topic proportions were related to author characteristics defined above. Third, we carried out a sentiment analysis regressing average response sentiment on the proportions devoted to each topic. The sentiment of a response was estimated using scores from the AFINN dictionary (Nielsen, 2011). This attaches a value between −5 (negative) to 5 (positive) value to words in the English language (e.g., admire has value 3, catastrophic has value −4). Note, given the free-text questions did not ask about government directly, some extracted topics were unrelated to our present purposes, though we report the results below.

Data cleaning and analysis was carried out by LW. AB selected government keywords. LW selected the number of model topics and LW and DF agreed upon narrative titles for the topics. A sensitivity analysis using unstemmed words produced similar topics.

## Results

### Descriptive statistics

Descriptive statistics are shown in Table 2. 13,298 (59.9%) individuals provided a free-text response, of which 4,402 used a government-related keyword (33.1%). 93.3% of those mentioning a government keyword provided a valid answer (e.g., containing a minimum of five words; n = 4,054). Supplementary Figure S2 shows the distribution of response lengths among those who used a government keyword, while Supplementary Table S2 shows the number of government-related responses by question. Most individuals used government related keywords in the question “What is bothering you the most about the pandemic? What aspects of it have you been finding most difficult?” (n = 2,701). More participants used government related keywords for the question on negative impacts on mental health and wellbeing (n = 384) than for the question on positive impacts on mental health and wellbeing (n = 69).

**Table 2:**
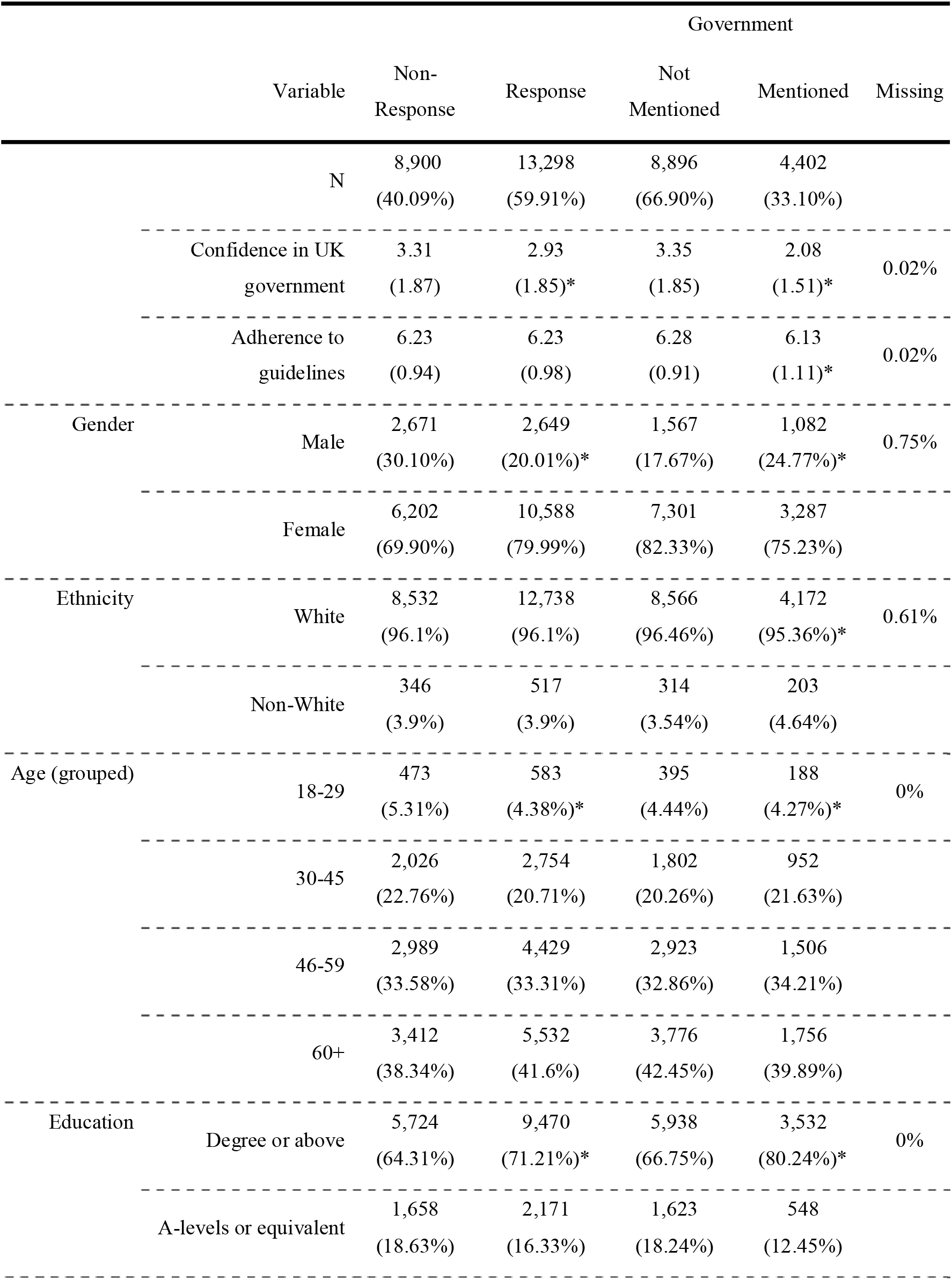
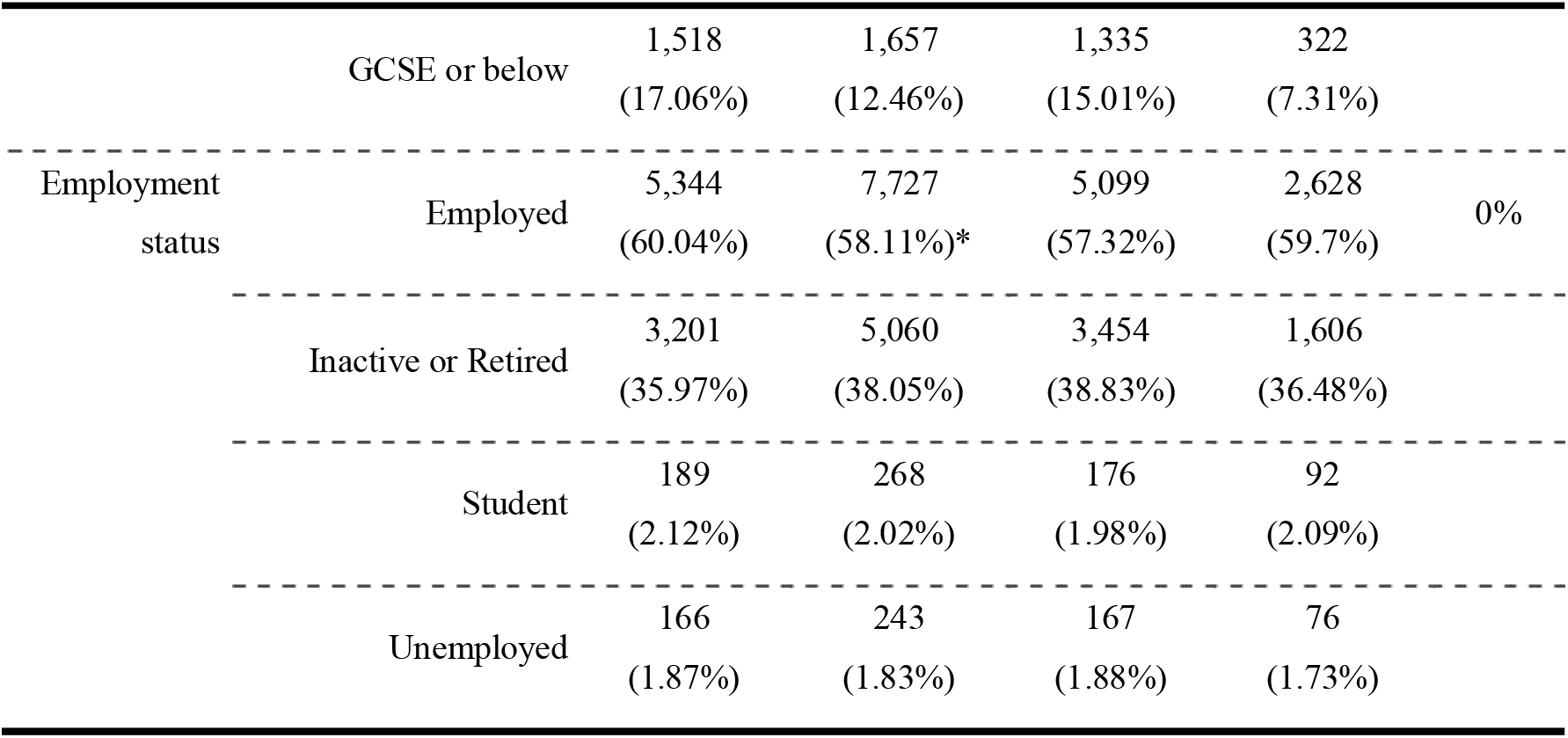
Sample descriptive statistics

Participants who provided a free-text response had lower confidence in government than non-responders on average, and were more likely to be female, older aged, and have degree-level education (Table 2). Similar patterns were observed among responders who used government keywords, specifically. Figure 2 shows the result of a multivariate Heckman selection probit model estimating the likelihood of responding and using a government keyword (conditional on responding) according to the person-level characteristics displayed in Table 2. Patterns are similar to those in the descriptive statistics. Given these responses biases – and the characteristics of the initial sample – our final sample is markedly different from the UK population.

**Figure 2.**
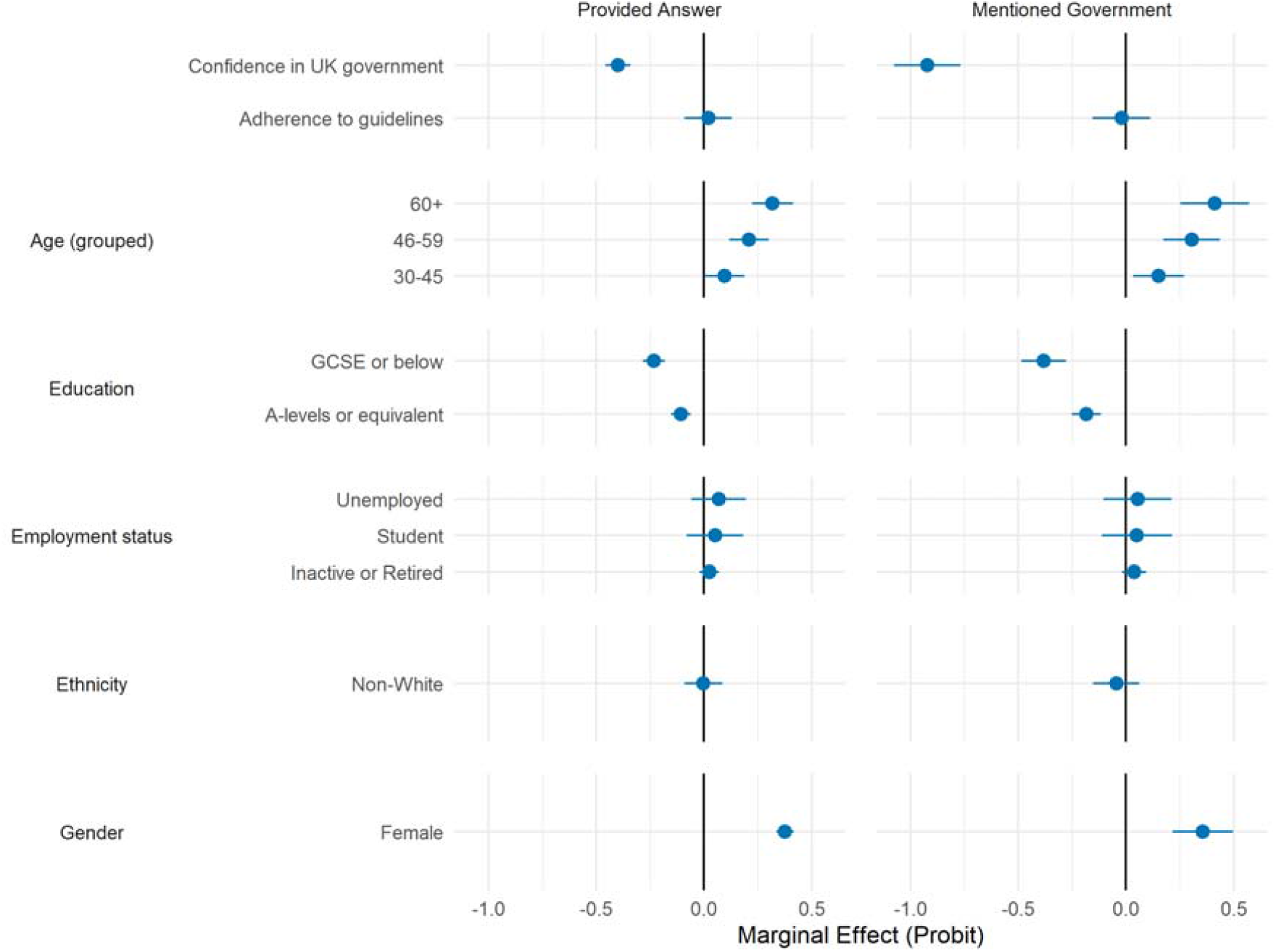
Marginal effect (+ 95% CIs) of participant characteristics and answering free-text questions (left panel) or mentioning government in free-text questions (right panel). Estimates derived from probit model (left panel) and Heckman’s selection probit model (right panel) with simultaneous adjustment for participant characteristics. Heckman’s selection model include each characteristic in both the selection and outcome parts of the model.

### Structural Topic Modelling

We identified an eleven topic solution. Topic descriptions are displayed in Table 3, along with short titles that we use when plotting results. These eleven topics clustered into three overarching themes containing 2-4 topics each (inconsistencies and uncertainties; tensions between government and others; disruptions to lives) and 3 standalone topics (see Figure 3), as follows:

**Table 3:**
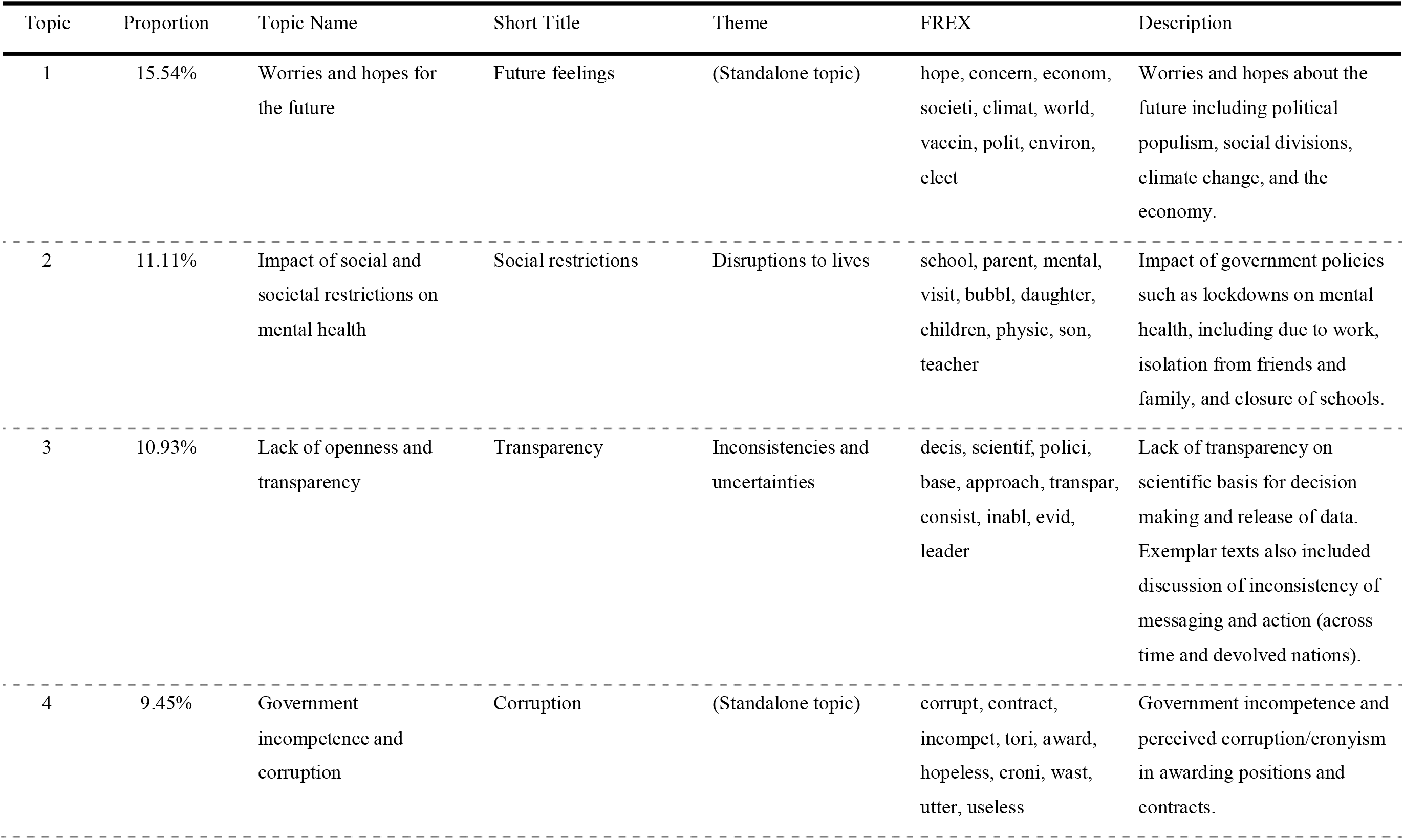
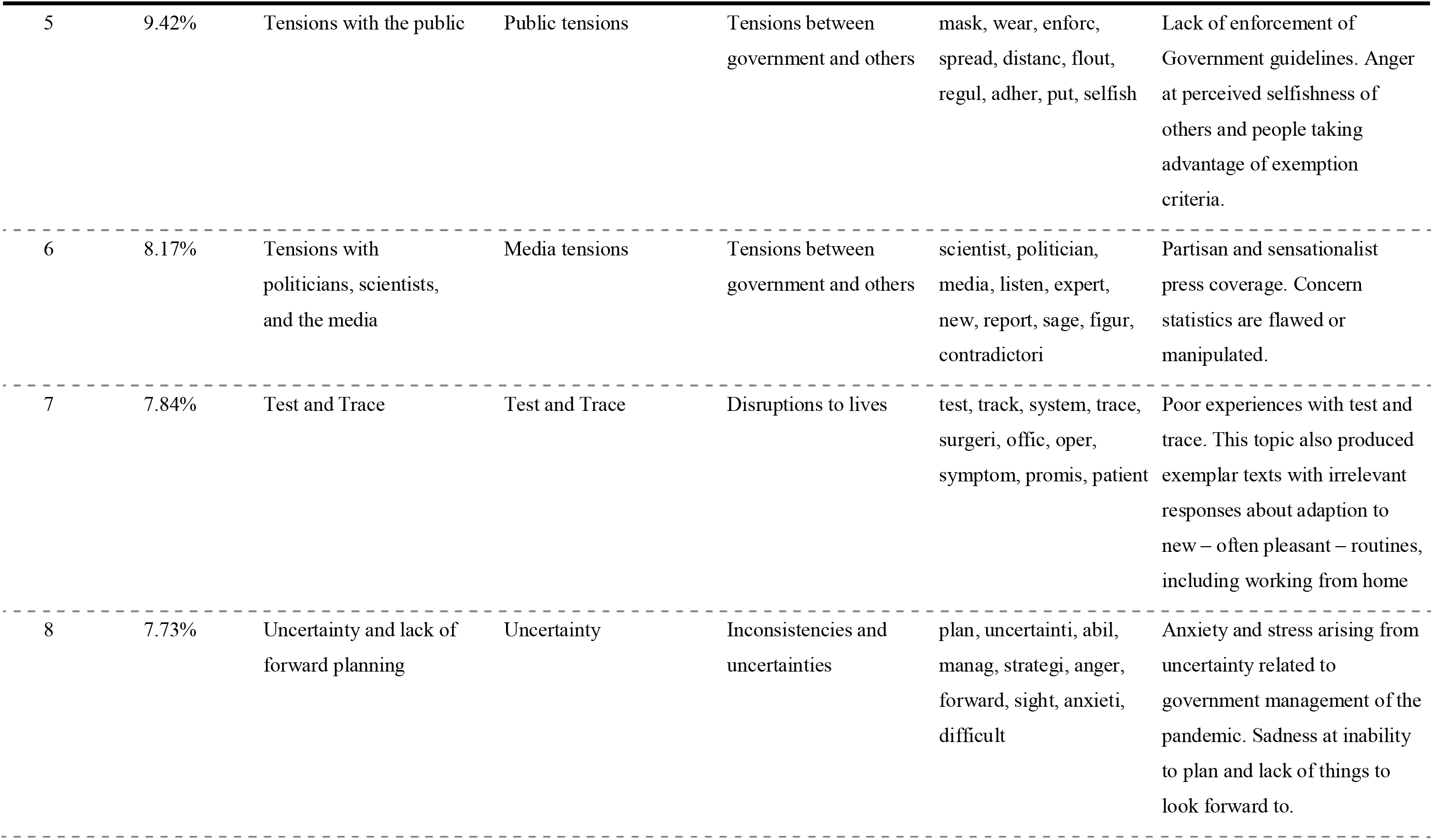
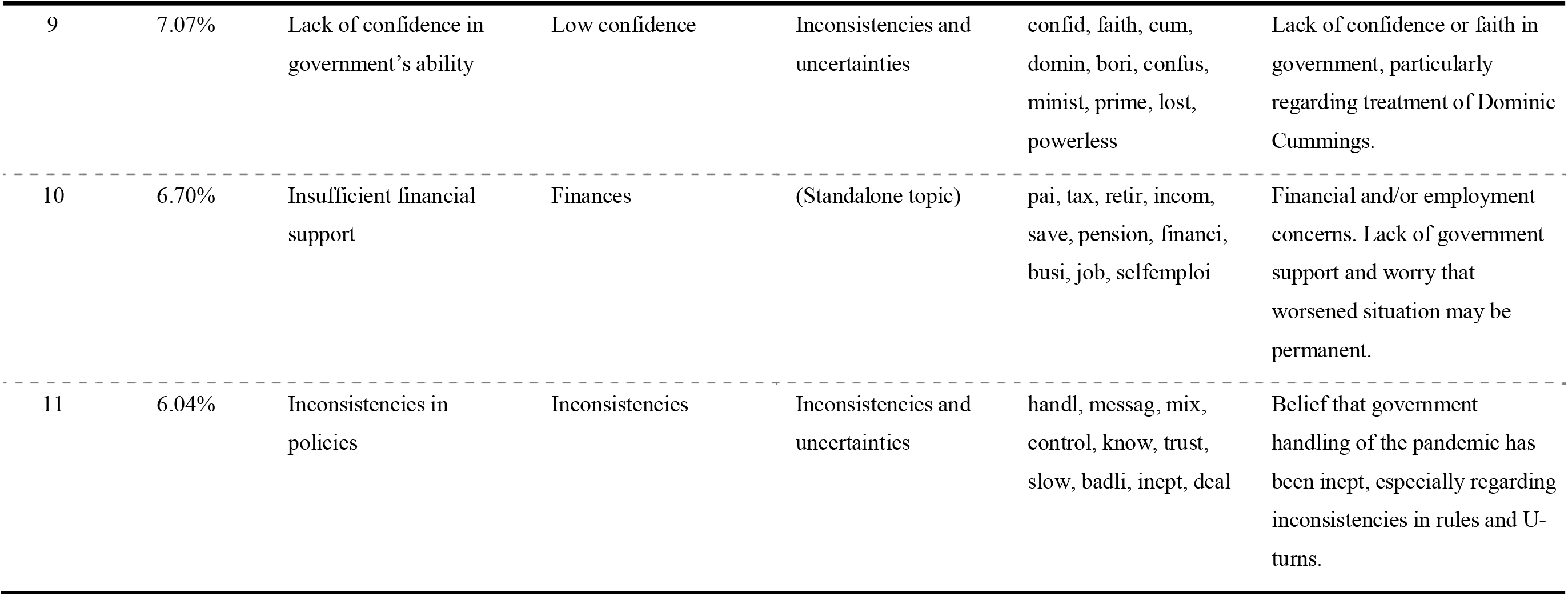
Topics extracted from structural topic model.

**Figure 3.**
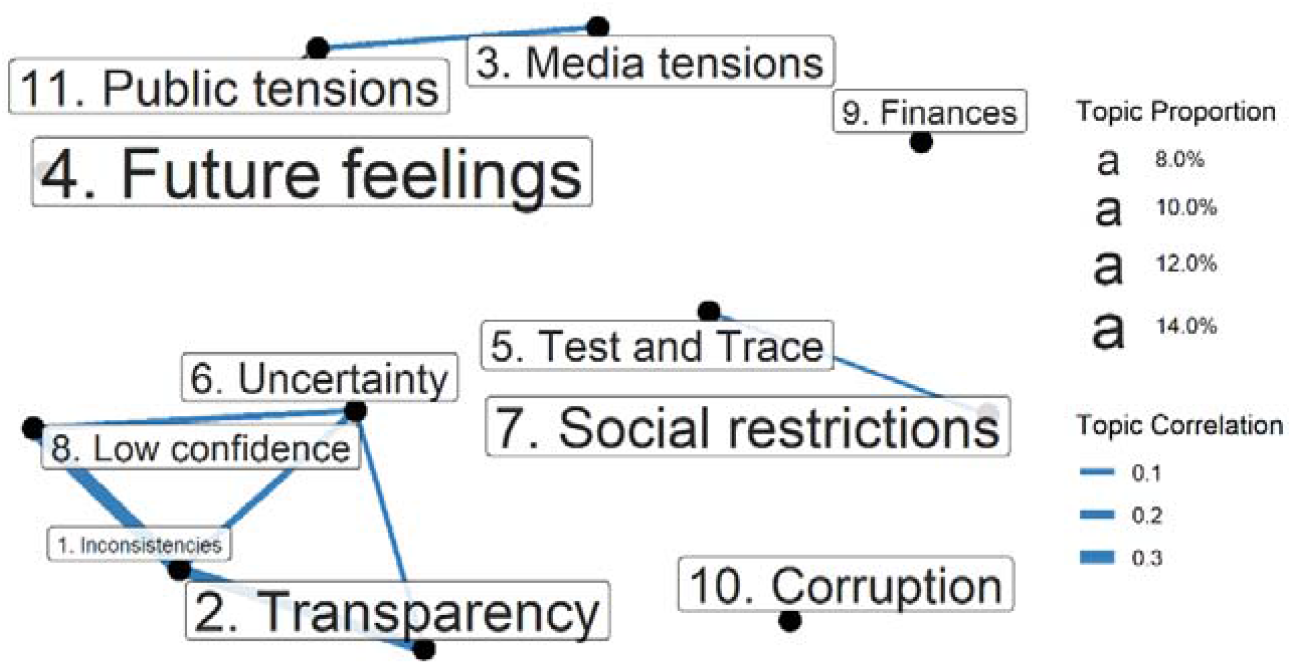
Correlation between topics. Correlations shown where ρ > 0.05

### Inconsistencies and uncertainties

The theme with the most topics focused on **inconsistencies and uncertainties** in the government’s actions.

### Lack of openness and transparency

The largest topic in this theme (topic 3; 10.9% of text) focused on **lack of openness and transparency** from the government over its decisions. Some expressed concern about the “*failure of the Government to govern well and to be transparent with the public*”. Examples included “*the absence of openness and transparency about the statistics being used by government to make major decisions*” and “*the refusal of government to present a balanced set of data analysing the pros and cons of lockdown*.” This was branded as “*unacceptable*” by some participants, who felt “*the public should be made aware of the balance of risk on both sides of the equation*.” It also made some feel that the government’s actions were “*illogical*” and that they were not following scientific advice, citing a “*lack of…scientific evidence from the Government explaining their decisions*” and beliefs that “*decisions are political rather than based on scientific evidence/advice*”.

This lack of transparency also led to challenges for members of the public to understand and follow rules and was seen as a deliberate tactic to control behaviours:

> *“It has been almost impossible for members of the public, without a great deal of digging into a number of different websites, to find out about the situation in their local area. I can only assume that this has been because the powers that be didn’t want the public to have access to local data because that might lead to them making their own decisions about risk in defiance of government guidelines.”*

These challenges were compounded by “*constant differences between the approach of the UK and devolved governments [and] a lack of cohesive effort*.”

Consequently, some participants reported that they would not follow guidelines in objection:

> *“The arbitrary nature of local restrictions and failure to give the public proper information in a partnership way is very frustrating. For this reason, I will not for example use the NHS app which has a nasty police state threat to it.”*

### Uncertainty and lack of forward planning

The second largest topic in this theme (topic 8; 7.7%) focused on the **uncertainty and lack of forward planning** from the government: *“there’s the understandable uncertainty of a pandemic -but that’s made much worse by the feeling that the Government doesn’t have a plan. It feels like there’s no way out.”*

Some participants felt that there had been little progress since the start of the pandemic: “*I feel that we are back to where [we] were in February/March this year 7-8 months later and that nothing has been properly implemented to mitigate the effects of the second wave*.” Some even expressed a “*lack of belief that [the] government has a plan*”, describing the planning as “*farcical*” and expressing “*anger with the government*.”

This uncertainty was perceived to have caused “*unnecessary stress and anxiety*” for participants:

> *“The ever changing restrictions have made me feel uncertain and anxious about what I can and can’t do. I am also frustrated at the lack of leadership and clarity from the government which has made me feel irritable and rebellious. The longer this situation goes on, the more anxious and irritable I feel.”*

This meant that some participants felt unable plan for the future: “*No coherent and effective strategy from central government robs us of any basis to confidently plan ahead*.”

For many, activities that usually helped to protect mental health had been disrupted (“*travel planning is usually a way of coping with depression for me*”) and “*not having things to look forward to*” was described as “*depressing*” and “*hard*”. This led some people to feel “*overwhelmed…I feel like I’m barely existing*.” As a result, some reported feeling a “*total lack of motivation to adhere to the rules*”.

### Inconsistencies in policies

A third topic in this theme (topic 11; 6.0%) focused on **inconsistencies in policies** during the pandemic. Many participants described the government’s response in strong language as “*shambolic*”, “*farcical*”, “*abysmal*” and “*piss poor*”: “*the UK government has handled it appallingly (even taking into account this is an unprecedented situation)*”.

Specifically, there were concerns about the “*lack of […] clear guidance*” and “*mixed messages*”. Some participants were concerned about “*U-turns by the government*” leading to a “*lack of control and…inconsistencies in rules [and] the apparent lack of logic to some of the government rules*.” Others spoke about the “*late response of central government*.”

Some felt that the government should have followed examples of other countries handling the pandemic more effectively: “*they have not looked at the example of countries that have been controlling it well, and instead have taken the approach of comparing our response to only those who are doing similarly badly*.”

Some were concerned that the policy inconsistencies meant that other people did not understand the rules they were supposed to be following or were not following them: “*While my household is very much sticking to the rules, I’m worried that other people won’t follow the rules*.” For some, they had followed the rules but without believing in them: “*Even though I have followed all their guidelines, I still find myself questioning everything they say and do*.”

### Lack of confidence in the government’s ability

Concerns about transparency, uncertainty and consistency were related to reports of a **lack of confidence in the government’s ability** (topic 9; 7.1%): *“the government seems to many amongst family and friends and myself incapable of charting a course that inspires us with confidence”*. Many participants expressed previously supporting the government, but this being eroded during the pandemic: “*I have lost all faith in the government*”. Specifically, some spoke about the government’s policies not seeming effective: “*the goverment [sic] seems to have no idea what to do and just keeps coming up with unworkable ideas*.” This was in contrast to people feeling that other governments had handled things better:

> *“It’s obvious to me that the Brits -and in fact most western nations are powerless to maintain the basic functions of government, to care for and protect the population… I’m going through the process of emigrating to Spain. I think I’ll feel safer in the EU for the time being.”*

Many specifically blamed “*the Dominic Cummings saga*”, labelling it as “*ridiculous*”. Participants reported that it had led to negative emotions: “*Since the Dominic Cummings incident, when it became clear that those of us who have obeyed not only the letter but also the intention of the rules were not considered the same as those with influence, the nation has become so divided and angry*.”

Specifically, people reported thinking that Cummings “*should have been sacked*”, explaining “*Boris sticking by him made everyone else’s sacrifices farsical [sic] and there was clearly a disregard for the rules after that. Shame on Boris and shame on Dominic*.” In the wake of the events surround Dominic Cummings, people expressed feeling foolish for having followed the rules themselves: “*I have kept to the rules. More fool me*.”

### Tensions between government and others

A second theme focused on **tensions between the government and others**, with others including the public, scientists and the media.

### Tensions with the public

A number of participants reported feeling a tension between the government and the public over the enforcement of rules (topic 5; 9.4%): “*The thing that bothers me the most about the pandemic is the fact that many people are not obeying the rules and are getting away from it*”.

Some directed their feelings at the public for not following the rules:

> *“It infuriates me that the wider public are ignoring the Government guidelines, the amount of people not wearing masks. The amount of ‘I’m exempt’ from wearing a mask*… *what exemptions? Illnesses? Then you should be staying at home to protect yourself surely?!”*

Specifically, some were frustrated that the actions of others affected their own ability to comply with the rules: “*You can do your utmost to protect yourself, but to some degree, you are still reliant on others socially distancing*.” Some proposed that the rules were not sufficient to contain the virus: “*Regardless of what announcements the government has made, the continuing spread of the virus and steadily increasing fatality rate show that their policies are being significantly ineffective*.” There were also calls to make the rules more stringent and to increase enforcement: “*I would like the government to be stricter and actually enforce the rules!*”. Some called for tighter rules on masks (“*the wearing of masks outside one’s home and garden should be made obligatory*”), others on lockdowns (“*the government taking so long to put us in lock down the first time, and for not long enough*”), and others on fines (“*The authorities and police should clamp down much more on those not following the rules and apply more draconian fines*.”).

But there were also some participants who disagreed with the provision of rules and questioned their true purpose:

> *“Bothering me most is the blatant lies being told by government and it’s so called experts. dont wear a mask they have no benefit wear a mask they save lives*….. *Sick of being told where I can go and who I can see… This is not about a virus it is about control and stripping freedom from citizens.”*

### Tensions with politicians, scientists, and the media

Other participants reported feeling a tension in the relationship amongst politicians and between government and both scientists and the media (topic 6; 8.2%).

There were expressions of frustration about internal tensions amongst MPs: “*[is it] too much to hope that politicians could work effectively and positively together to help in the outcome of the virus, instead they continue to bicker and play the blame game, whilst in some cases, not even being good role models*.” People were concerned that the pandemic had “*all become very political*”.

Some felt that the government was not listening to scientists: “*it seems as if Borris [sic] Johnson will not listen to Medical advice and wants to do what he wants*.” But others felt that the government were being led too much by the scientists, expressing concern about “*what the scientists are advising the Government to do*”, with concerns that the “*statistics are flawed*” and governments were “*giving over-exaggerated information about the true effects of Covid-19*” and not doing “*the right thing*”. This was all underpinned by concerns from participants about which facts were “*the true facts*”.

A number were critical of the media for highlighting these tensions: “*The public does not need to know about the wranglings between experts and politicians, just the expert advice which impacts our lives*.” There were even concerns that the media was undermining, or even pursuing agendas against, the government: “*Anger. At the appalling tv news coverage of all 3 main stations. Their sole aim is to try to embarrass, interrupt and bring down government representatives… Appalling*.” But others felt that the media should be applying more scrutiny to what the government said: “*Main Stream Media not doing their job…[they] need to Report FACT and apply Investigative Journalism, not just repeating press releases and giving out the wrong information*”

The media was also criticised for other aspects of its coverage. Some felt that media coverage had been “*scaremongering and inaccurate*”. This led a number of participants to report that their mental health had been affected by the “*sensationalised headlines*”.

### Disruptions to lives

A third theme was around the “*disruptions to routines*” brought about by government policies and how these had affected day-to-day lives and mental health.

#### Impact of social and societal restrictions on mental health

The second largest topic to emerge in this study was about policies relating to social & societal restrictions. This topic was presented strongly in terms of how such restrictions affected people’s mental health as a result of their impact on work and family life (topic 2; 11.1%).

Many described how the lockdown measures had put enormous strain on their families. Some described parenting challenges (“*my relationship with my 6 year old boy, only child, has deteriorated because we are constantly fighting over screen time limits*”), others disruption for children being home-schooled (“*[my daughter] was diagnosed with anxiety due to lockdown. All she wanted was for life to be normal*”). Some felt the government had not considered the impact on children enough:

> *“I feel so strongly that the Governments [sic] stance was very much that young children are not affected by Covid-19 and would not fall ill or die from it. However, the impact it has had on their mental health by not being at school…has been so sad…I feel so strongly that they are the lost generation through all of this and this will impact our society for many years to come.”*

However, others expressed how there were now additional pressures when lockdown measures eased (“*I drive son to/from college everyday due to risk of covid on bus…a 45 minute trip…twice a day*”) and less time as parents had to return to work whilst still coping with the disruptions of the pandemic (“*during lockdown i [sic] was able to do Joe Wicks and walk everyday, now I don’t have the time or energy*”). People felt that these family pressures were not being considered in planning policies around social restrictions: “*What the government are doing to families is cruel, unfair and unjust. Their only concern now is the economy*.”

Finally, many reported being upset by the rules limiting contact with friends and relatives, with common phrases focusing on “*missing hugs*”. Some were upset that they could not have contact with others in nursing homes (“*My sister lives in a nursing home, i would normally visit 4 times a week. She hasnt [sic] been allowed visitors since March. She is now not allowed to leave the home and is virtually being kept a prisoner*.”) They felt that the rules were unfair:“*people can go to pubs restaurants and shops but you cant visit elderly of [sic] sick relatives*” Others felt that support bubbles rulings were “*very unfair*”, either because they were not allowed to form a bubble or because they had “*no one to form one with*”.

The restrictions imposed within society also caused other concerns, including “*worry about finances*” and employment, with some expressing “*frustration at the lack of government understanding of vocation*” in thinking that people could be furloughed indefinitely without it impacting their work skills. Specifically, some expressed concern that the government had overlooked certain groups and their needs (for example, “*the government have not considered our armed forces living abroad*”). Many keyworkers were upset that the challenges they faced had not been properly considered within social restriction policies. For example, teachers raised concerns about parents sending “*children into school poorly because of financial implications of having to isolate*”, putting them at risk, whilst social care workers reported disruption in their jobs: “*my job in adult social care…has been changed twice since March, and i am under more pressure at work and worried about losing my job due to cuts*.” This led to reports of keyworkers feeling underappreciated: “*My life is not valued by the government!*”

There were also anxieties relating to accessing health services, with many feeling that the government had not provided enough support to the NHS during the pandemic leading to restrictions in what support people could access. Reported challenges included accessing mental health care *(“[there is] little access to mental health support [my daughter] needs*”), not receiving cancer treatment (“*my partner is not getting treatment because he is high risk and vulnerable, so they don’t want him travelling to hospitals*”), not seeking medical help (“*because they are scared of dying in hospital alone*”), and not having support through maternity care (“*anxiety about restrictions on my partner being present during labour*”).

Many reported that these experiences of difficulty created by the rules and their effects on mental health put them in an awkward position in complying with the rules: “*It’s very frustrating and you often feel torn between following government instruction and protecting your own mental health*”.

### Test and Trace

Another topic to emerge specifically covered people’s experiences of Test and Trace and the impact it had on their lives (topic 7; 7.8%).

Many described challenges specifically relating to test and trace: “*my experience of test and trace has shown total incompetence*”. People reported finding the app unreliable (“*my husband…tested positive…but was not contacted by test and trace until a week later*”), inconsistent (“*there seems to be no consistency in the advice compared to the government’s website advice*”), and untrustworthy:

> *“I was told to self-isolate for nine days -why nine days? By the NHS a tracker app. Very disconcerting, NO info about where/when I might have been in contact with someone infected. Very frustrating…was dubious about whether it was valid.”*

Others had poor experiences in being over-contacted (“*we have been told there is no record that the calls are being duplicated and in fact I was made to feel like I was actually lying*”) and treated badly (“*I was very angry at our family’s treatment by test and trace [but] there was no clear way to complain*”).

Some felt the government had not considered the impact of test and trace on their work: “*[I had] to cancel 2 weeks [sic] worth of clients to isolate after a client showed symptoms, despite wearing the correct PPE and following hygiene guidelines*.” Specifically, losing out financially due to having to isolate was a common complaint: “*because we’re not on benefits we couldn’t claim any help other than SSP, this is £500+ per week that we’ve lost*”. This led to some people feeling “*undervalued by the government*”.

Overall, many concluded that “*the test and track system is not fit for purpose*” and reported not wanting to use it: “*Leaves me tempted to turn the tracker off*”.

### Insufficient financial support

A further topic raised by participants focused on **insufficient financial and employment support** provided by the government during the pandemic (topic 10; 6.7%).

Although participants acknowledged the government support schemes, many participants reported such measures not reaching everyone equally nor going far enough. Difficulties finding work amongst those who had lost jobs were common: “*with an increase in unemployment there are more people for less [sic] jobs*”. Many people also described substantial financial problems, often due to loss of work. A number reported they had “*slipped through the net*” to qualify for financial support, for example because they had only recently set up their own business (often involving “*considerable outlay prior to lockdown*”). There was a perceived injustice amongst many of these people that as taxpayers they were “*expected to share the burden of the cost” of such schemes yet had “not received a single penny of this assistance and support*”, with some describing it as “*fundamentally and inherently wrong*”.

Even those with savings were concerned as savings that were intended “*for maybe a wedding a home, retirement or kids [sic] education*” meant they were ineligible for current government support. They worried about the long-term impact of “*running out of savings as it is all i have to last me to my pension and the Gov keeps moving that further away*”. Many felt they would never “*be able to recoup my costs*”. Some had had their savings “*decimated [through] the falling stock market*”. This was particularly a cause for concern amongst people who were retired and “*dependent on income from my savings and investments to survive*”.

As a result of these challenges, some perceived themselves as being “*totally overlooked and forgotten by this government*” and “*completely shafted*”: “*I don’t understand what the government is playing at destroying peoples businesses and livelihoods running up enormous debt and destroying the economy*.” This was described as “*the government’s ‘couldn’t care less’ attitude*” and “*‘head in the sand’ attitude*” and labelled “*pathetic*”. There was also anger at the inequalities being created by the pandemic: “*Some people getting richer doing nothing, some people getting nothing and suffering*.”

Many described detrimental effects on their mental health as a result of insufficient financial support, including “*causing me very significant stress, anger and resentment and is having an enormously detrimental effect on my mental health and wellbeing*.” Some went so far as to describe themselves as “*desperate*” and referred to people they knew who had “*taken their own lives*.” There was a concern that the government had “*put money ahead of the health of the people… ‘What are a few lifes [sic] compared with the business losses!*’”

### Government incompetence & corruption

Another topic to emerge focused on perceived **government incompetence and corruption** (topic 4; 9.5%).

Many participants described a perceived unfairness in the way the government was providing contracts for work during the pandemic. Many were concerned that the government was “*bypassing NHS and giving out expensive contracts that belong within NHS to private companies owned by their donors leaving NHS without appropriate funding*”, describing it as “*corruption plain and simple*”. There was also concern that the government had “*defunded and hamstrung the NHS and Social Services*” in the lead up to the pandemic, leaving it unable to operate as well as it could have done.

Others felt that the government had been “*using the pandemic as a cover for handing out huge enormous contracts*” to “*friends of the government*” who were “*inexperienced and unsuitable*” to carry out the work, leading to “*no prospect of receiving services (e*.*g. the multi-million contract for PPE that couldn’t be used).”* These actions were felt to be “*greedy*” and “*self-serving*” of the government and “*examples of cronyism*” and “*complete incompetence and total corruption*” and were reported with high degrees of emotion (“*it has bothered me endlessly*”, “*feeling stressed and angry*”, and “*it is hugely depressing and infuriating*”).

Some were specifically concerned that the costs of contracts awarded during the pandemic were “*wast[ing] vast sums of public money*” and taking it away from other priorities such as “*the huge increase in poverty and child hunger which this joke of a government is failing to take seriously or address*”.

### Worries and hopes for the future

Finally, the largest topic (which did not come under any broader themes but stood alone) was around people’s **worries and hopes for the future** (topic 1; 15.5%).

Participants reported being “*dispirited*” that the pandemic was “*an inevitable consequence of human interventions”*. Some were worried about threats directly as a result of the pandemic:

> *“Fear of what instability the future will bring because of the posdible [sic] collapse of the economy/the government /food supply/ medical supplies. Civil unrest could happen, wars could increase. The infra structures we rely on could crumble or be dismantled. Chaos could bring power to really evil people. Or a more subtle and nuanced version of any of the above.”*

Others were concerned that the pandemic had detracted from important political agendas: “*Concerned, the pandemic struck at a time when society, the country and the world face challenging times (e*.*g. Brexit, rising populism, national and international divisions, environmental challenges etc*.*). I tend to optimism, but Covid 19 is a disruptive factor that detracts from tackling these other pressing matters*.” Specifically, there were worries that “*Covid-19 will exacerbate existing social and economic divisions*” and that “*the momentum for dealing with climate change will be lost with increase in single use plastic use and car journeys favoured over public transport etc*”. Brexit was a common concern alongside Covid-19 (“*Concern about Brexit being even more damaging that [sic] it was pre-covid*”).

Some were concerned that they would not see a recovery from the ill effects of the pandemic (“*sadly, I am not hopeful that any of that will be reversed in my life time*”), with some describing themselves as “*pessimistic about the long term world economy, global political unrest, further world health issues, and environmental degradation*.” There were also participants who felt more keenly aware of the future threat of pandemics as a result of Covid-19: “*I am worried that even after covid 19 we will see more & more pandemics & it will effect my children’s future*.”

However, others expressed optimism. There was appreciation for “*kindness and humanity at its best [being] demonstrated*” during the pandemic and some felt “*positive about rethinking what is really important in life both on a personal level and for societies*”. This hope included “*that people will value slower pace, time outside and friends over more and more unsustainable consumption of resources*”.

### Topic proportions and author characteristics

In exploring how topic proportions varied by participant characteristics, a number of predictors emerged (Figure 4). Topics 3 (transparency), 4 (corruption), 8 (uncertainty), 9 (low confidence) and 11 (inconsistencies) were expressed more commonly amongst people who reported lower confidence in government to handle the pandemic, while Topics 5 (media tensions), 6 (public tensions) and 10 (finances) were expressed more commonly amongst people with higher confidence in government. Regarding compliance with guidelines, individuals perceiving that media coverage has been sensationalist (Topic 6) and reporting greater financial issues (Topic 10) were more likely to report lower compliance. The evidence that expressing anger at other’s non-compliance (Topic 5) was related to personal compliance was weak. Women were less likely to discuss government corruption (Topic 4), transparency (Topic 3), and issues with the media (Topic 5), but were more likely to discuss problems from social restrictions (Topic 2). There was little difference by ethnicity. Older individuals were more likely to write about government corruption (Topic 4), while younger people were more likely to note others’ non-compliance (Topic 5), as were individuals with GCSE or lower education. Unemployed individuals were more likely to discuss financial and employment difficulties (Topic 10).

**Figure 4.**
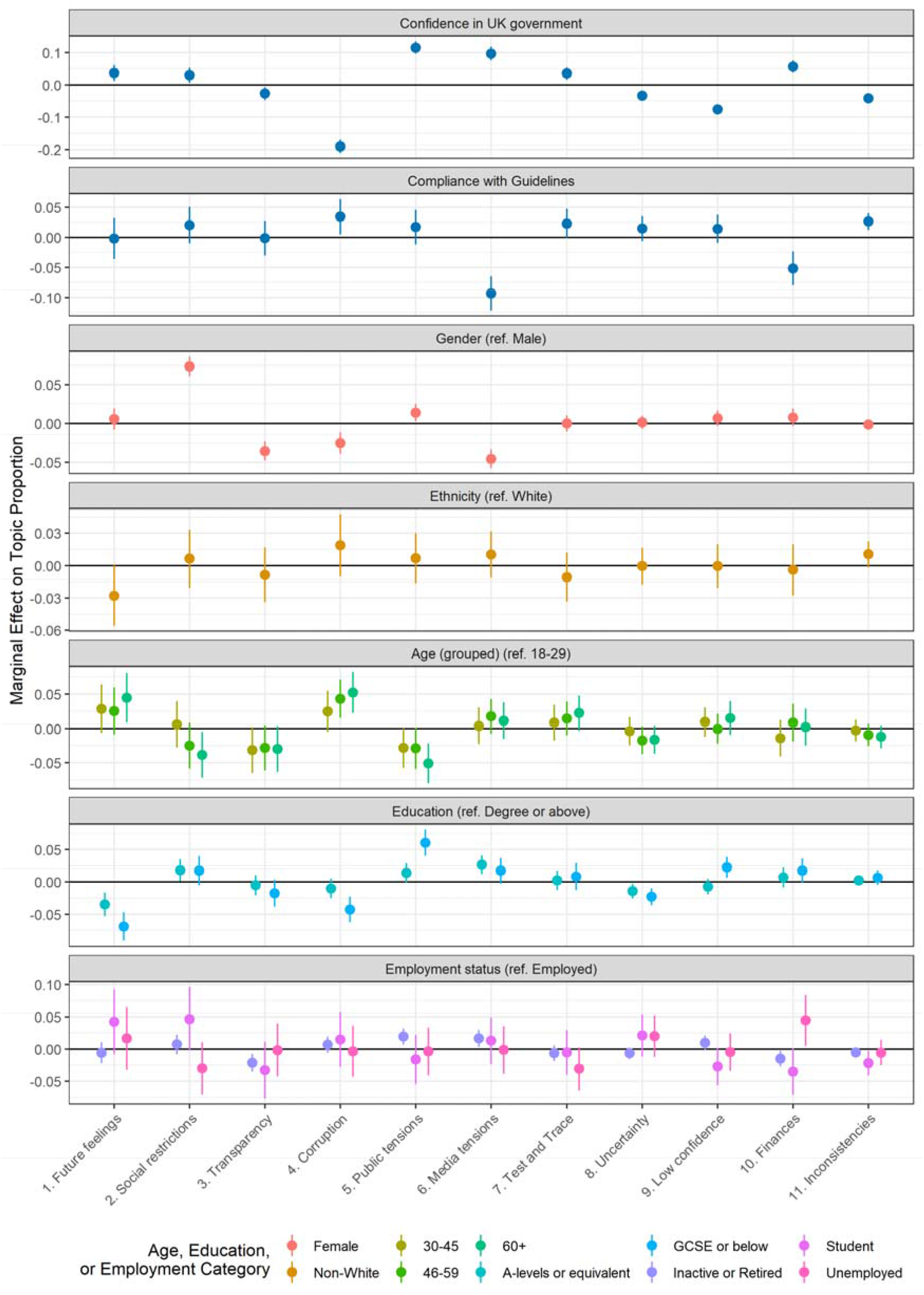
Association between topic proportions and person characteristics (+ 95% CIs). Drawn from multivariate regression models. Continuous variables scaled to 0-1 range, so estimate reflects estimated change in topic proportion due to increase from minimum to maximum value.

### Sentiment analysis

To identify how positive or negative responses were overall, we carried out sentiment analysis. Figure 5 displays the results of a multivariate regression assessing the association between the overall sentiment of a response and topic proportions. Topic proportions for each topic were added to the model simultaneously. Eight of the 11 topics were clearly related to more negative sentiments, with topics 11 (inconsistencies) and 4 (corruption) showing the greatest level of negative sentiment. Respondents in topics 1 (future feelings), 7 (Test and Trace) and 9 (low confidence) were related to responses that were more neutral, though this is llikely to be partly due to high probability words not having sentiment (e.g., Bori[s], Domin[ic]).

**Figure 5.**
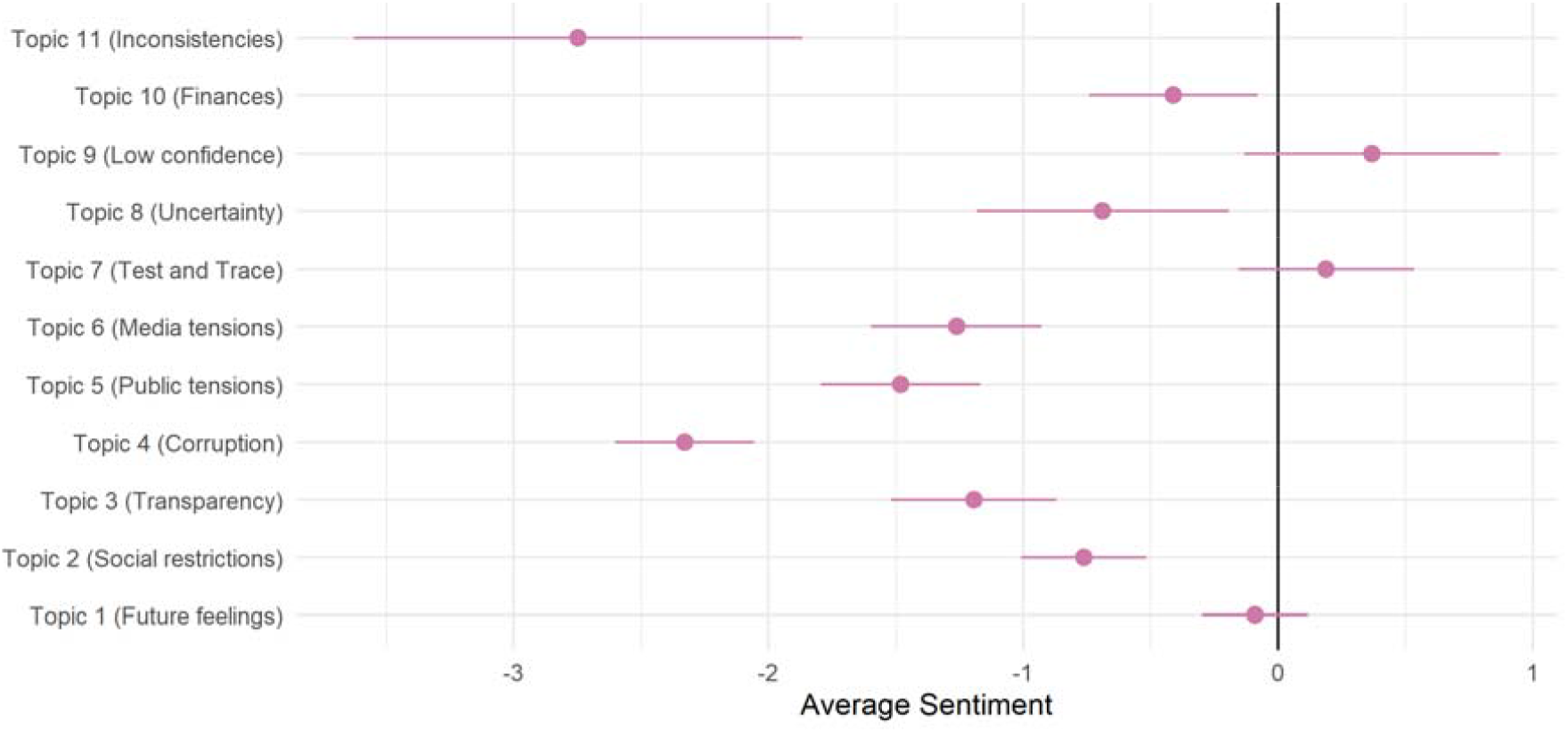
Sentiment analysis. Results of linear regression of average sentiment on topic proportions in text.

## Discussion

Using free-text data from 4,000 adults in England, we identified eleven topics that highlight the concerns of the public regarding the UK government’s handling of the COVID-19 pandemic. Five of the topics related to beliefs about government incompetence, lack of planning, inconsistency in policy decisions, and allegations of government cronyism and corruption. Participants also wrote about difficulties with the Test and Trace system and insufficient support protecting from financial losses due to the pandemic, for instance, resulting from individuals not meeting eligibility criteria. Individuals who were more supportive of the government were more likely to offer criticisms of non-government figures, such as the public for violating guidelines and the media for sensationalist coverage. Some of the topics identified were clearly related to participant characteristics, with notable age differences in the topics raised. Most concerningly, the responses showed public health implications of the government’s response to the pandemic. Participants repeatedly reported difficulty keeping up with current guidance, demotivation from complying with the rules and even deliberately not following the rules in reaction to government actions, and described impacts of government policies and behaviours on their mental health.

These results add detail to previous survey data showing low confidence in the UK government across the pandemic (Supplementary Figure S1; YouGov, 2021). Although free-text questions asked participants about both positive and negative feelings, and experiences and responses were not clearly related to political persuasions as there were examples in our data of people disclosing that they were Conservative voters and expressing dismay at the government’s actions (see also Opinium, 2020), our sample was not nationally representative. Participants who chose to answer free-text questions and chose to mention the government in their responses had on average slightly lower scores on scales measuring their confidence in the government than the rest of the sample. Therefore, this study does not claim to be a study of all political opinions during the pandemic, but rather aims to show how concerns about the government’s handling of the pandemic manifested themselves, how they were related to one another, and what their implications were for public health.

Some of the topics identified in this study echo findings from prior qualitative studies carried out during the COVID-19 pandemic. Studies using data from convenience samples (Williams et al., 2020a, 2020b) or specific groups, such as BAME and low-income groups (Denford et al., 2020), health and social care workers (Aughterson et al., 2021; Hoernke et al., 2021) and individuals with respiratory illnesses (Philip et al., 2020), have identified themes related to poor government management of the pandemic, including lack of clarity in communication, inconsistent guidance, and uncertainty due to lack of government planning. Many have complained that members of the public have not been complying with guidelines and that the government should enforce rules more strictly (Aughterson et al., 2021; Philip et al., 2020; Williams et al., 2020a). It is further notable that a number of participants spoke negatively about the political response to the news that Dominic Cummings broke lockdown rules, consistent with previous evidence that confidence fell following initial reporting of this incident (Fancourt, Steptoe, et al., 2020). Given the data in this study were collected almost six months later, the results suggest the effects on confidence have been lasting. But our results build on these previous findings by showing how these concerns co-occur and are related to participant characteristics and that these may be concerns in the population at large.

Specifically, the evidence here suggests that aspects of the government’s response to the pandemic may be undermining efforts to tackle the virus. Participants noted that inconsistent and unclear messaging had made it difficult to keep track of current guidelines, with some reporting inadvertent violations of rules. This finding supports those from other recent studies that have shown a relationship between unclear messaging and individuals interpreting rules in a favourable way (Denford et al., 2020; Williams et al., 2020a, 2020b), as well as frequent changes in the rules leading to “alert fatigue” and participants not taking in latest advice (Williams et al., 2020b). Whilst changes to the rules may be necessary to be responsive to risk (Denford et al., 2020), there are important implications to consider in terms of how this affects compliance.

Another public health consequence of the government’s actions during the pandemic was reported; adverse effects on mental health. Whilst mental health has been shown to be affected during the pandemic in a number of previous studies, much of the focus has been on causes such as social isolation, lockdowns, disruption to healthy behaviours such as exercise, and anxiety about the virus. Our findings on the impact of social restrictions on mental health support these previous findings, but they also highlight the effects of other behaviours and government policies. In particular, uncertainty arising from perceptions of a lack of government planning caused stress and anxiety and stopped individuals from making future plans; a theme found in a prior qualitative study (Philip et al., 2020). Further, a lack of financial support for all individuals affected by the crisis also impacted mental health, in line with existing quantitative studies showing that financial stress was related to worsened mental health during the pandemic (Chandola et al., 2020; Wright et al., 2021).

This study had a number of limitations. First, we did not use data from a representative sample of the UK population and individuals who chose to respond and who spoke about the Government were a further sub-sample. However, the sample was heterogeneous and all participants were given the option to provide free text responses. The questions asked did not directly refer to opinions on the government, which would have had the limitation of potentially prompting participants to take particular sides, so the opinions that were given were salient and held strongly enough that they were offered without direct prompt. Further, our focus was not on polling all opinions on the government but rather on understanding patterns and consequences of concerns about the government’s handling of the pandemic. It is also notable that the sample who responded was not dominated by those who were unemployed or from minority groups most adversely affected by the pandemic, whose experiences may be expected to have been the worst. Second, we identified government-related responses with government keywords but in some cases, these keywords were homonyms not necessarily used in a government-related context – for instance, “labour” was sometimes used to refer to childbirth instead of the political party. Further, as responses were not constricted to be about the government, some text was unrelated but was still included in the analysis, leading to topics that were not always fully homogeneous, so associations in the regression modelling and sentiment analyses may be driven by a combination of relevant and non-representative texts. However, the structural topic modelling provided a good solution and the results are in line with previous research. Third, while we used a large set of government keywords, these were not exhaustive. For instance, we did not include keywords relating to specific policies such as lockdown or furlough. However, participants raised such policies in their responses.

Nevertheless, this study provides detailed insight into the views of adults living in England on the UK government during the COVID-19 pandemic. It shows public concerns about inconsistencies and uncertainties during the pandemic, tensions between the government and the public and media, disruptions to individuals’ lives as a result of government policies and programmes, worries about government incompetence and corruption, and the implications of financial policies. It also highlights some of the most pressing concerns that individuals have as the pandemic continues and beyond, and suggests that policies are needed that enable people to safely maintain social connectedness as well as provide financial support to those whose work has been affected. This, alongside consistent and transparent communication and messaging from the government, is critical to improving compliance with measures to contain the virus, as well as protecting mental health during health emergencies.

## Data Availability

The code used is available at https://osf.io/jw3gb/. The data are not available due to stipulations set out by the ethics committee.

https://github.com/UCL-BSH/CSSUserGuide

