## Supplemental Information for "Public Opinion about the UK Government during COVID-19 and Implications for Public Health: A Topic Modelling Analysis of Open-Ended Survey Response Data"

Supplementary Information

### Political Context

The period around 14 October to 26 November overlapped with several key developments in the UK Government’s response to the COVID-19 pandemic in England. On 12 October, Boris Johnson announced a three-tier system of regional restrictions taking effect from 14 October. One area, Liverpool, was assigned the strictest restrictions. On 22 October, increased financial support for jobs and workers was unveiled, with the package announced shortly after areas of the south of England were placed into higher tiers. On 27 October, the UK recorded 367 deaths from COVID-19, the highest daily total since May. Four days later, the government announced a second four-week national lockdown in England, coming into effect from 5 November. Retail and leisure venues closed and individuals were allowed to meet with at most one member of another household in an outdoor setting. The government’s job furlough scheme was extended to the end of March 2021. On 9 November, Pfizer and BioNTech reported effectiveness of 90% in human trials of their COVID-19 vaccine. On 18 November, the National Audit Office released a report finding that politically connected suppliers of PPE were 10 times more likely to be awarded contracts during the pandemic (National Audit Office, 2020), and on 21 November, campaigners announced their intention to take legal action against the UK government over the appointment of politically connected individuals to key roles for tackling COVID-19 (Good Law Project, 2020). On 26 November, a new tier system was announced for England, with news that many parts of the Midlands and the North of England would be placed in the highest tier when the lockdown ended.

### Data Cleaning

We performed topic modelling using unigrams (single words). Responses were cleaned using an iterative process. The main steps were as follows. Popular hyphenated words were collapsed into non-hyphenated form and spaces were removed between words that could have been hyphenated (e.g., “pre-pandemic” and “pre pandemic” became “prepandemic”). Punctuation mistakes (e.g., full stops between words) were replaced with whitespace unless the full stop denoted an initialism or a URL. Full stops were removed between initialisms - e.g., U.K. became UK - and “www.” was removed from URLs. Spelling mistakes made in 10 or more instances across responses were corrected manually with spelling mistakes were identified using the Hunspell algorithm (Ooms, 2018). All misspellings of “government”, “government’s” or “governments” were corrected. Accented characters were replaced, contractions expanded (e.g., “hasn’t” to “has not”), and possessive forms (trailing “‘s”) removed. Responses were tokenized into lower-case unigram form and “stop” words (common words such as “the” and “and”) were removed. Stop words were identified with the onix (Lextek International, 2021), SMART (Lewis et al., 2004), and snowball (M. Porter & Boulton, 2002) dictionaries. We excluded the stop words “work”, “working”, “worked”, “works”, and “help” given their relevance to the current topic. To reduce data sparsity, in the STM analysis, we further stemmed words using the Porter (1980) algorithm, dropped responses if they contained fewer than ten words, and dropped words if they appeared in fewer than five responses (Banks et al., 2018).

Data cleaning was carried out in R version 3.6.3 (R Core Team, 2020) using the tidyverse (Wickham et al., 2019), stringi (Gagolewski, 2020), qdap (Rinker, 2020), hunspell (Ooms, 2018), SnowballC (Bouchet-Valat, 2020), and tidytext (Silge & Robinson, 2016) packages. The code used is available at <https://osf.io/jw3gb/>. The free-text data are not available due to stipulations set out by the ethics committee.

### Free-Text Modules Questions

Question

*Q1. Is there anything you would like to tell us about the changes that have been brought about by the Covid-19 pandemic and the impact these have had on your mental health or wellbeing?

*Q2. What is bothering you the most about the pandemic? What aspects of it have you been finding most difficult?

*Q3. Has the pandemic had any negative impacts on your mental health and wellbeing? If so could you tell us about these?

*Q4. Has the pandemic had any positive impacts on your mental health and wellbeing? If so could you tell us about these?

Q5. How have your circumstances (e.g. work, housing, local area, finances, social networks, family life, responsibilities etc ) contributed to your experiences (positive, negative or both) of the pandemic?

Q6. How have your personal attributes (e.g. age, gender, ethnicity, sexuality, health conditions etc) contributed to your experiences (positive, negative or both) of the pandemic?

Q7. What have been your methods for coping during the pandemic so far and which have been the most or least helpful?

*Q8. Since the Covid-19 pandemic began, how have you been feeling about the future? What are you hopeful or concerned about?

* Question used in this analysis

### Figures

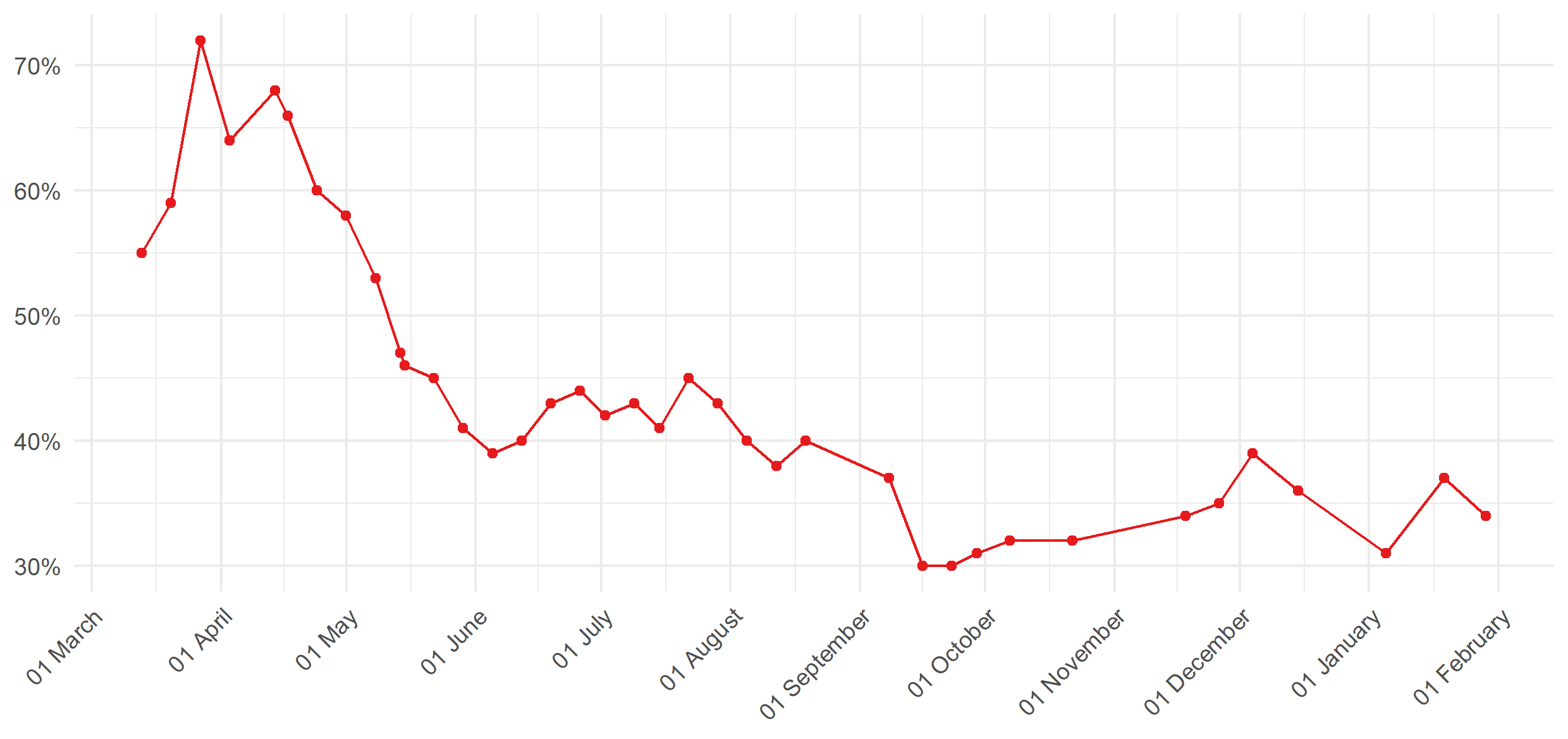

Figure S1: Proportion of respondents stating that government in handling COVID-19 “very” or “somewhat” well. Source: YouGov (2020)

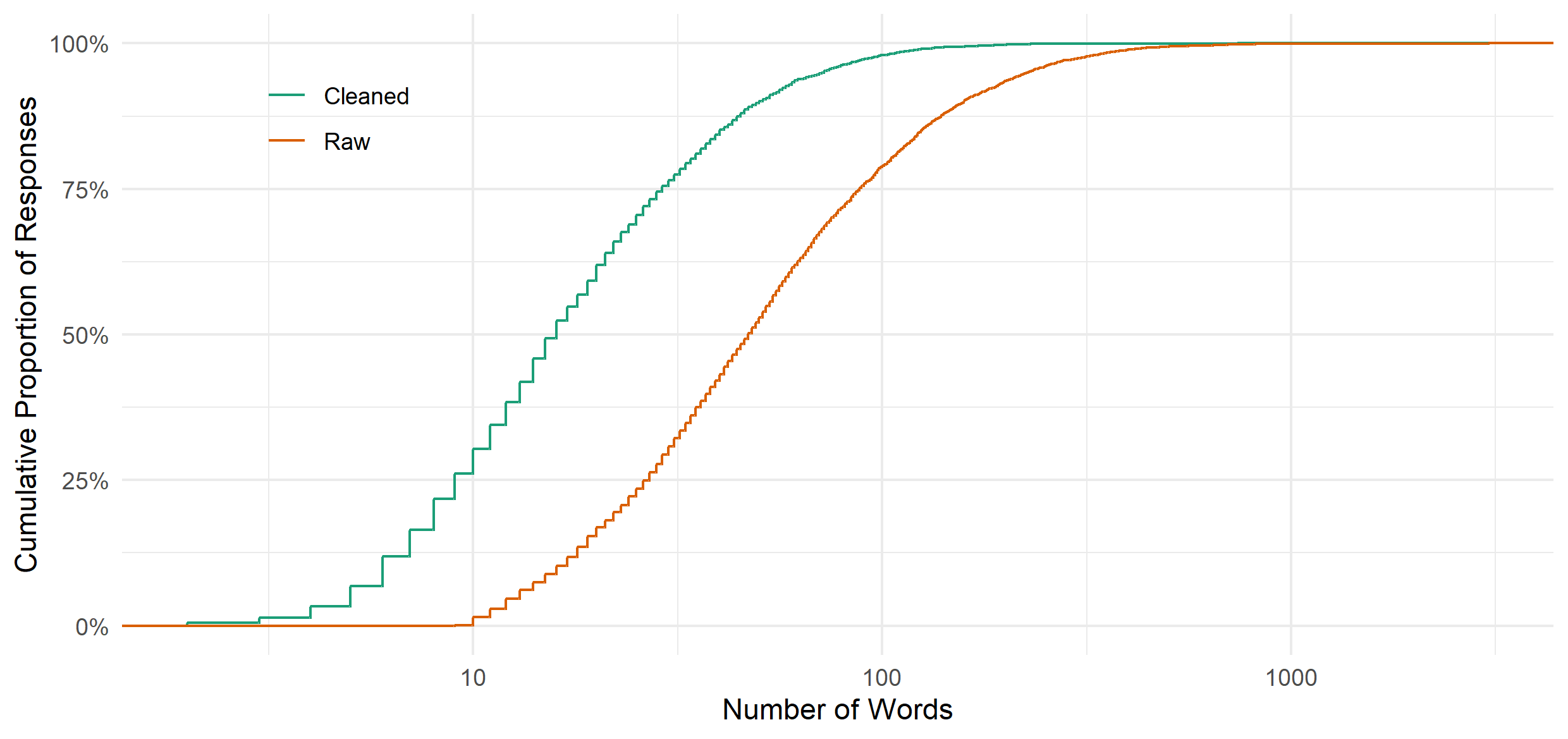

Figure S2: Cumulative density curves for number of responses by number of words in (cleaned or raw) document.

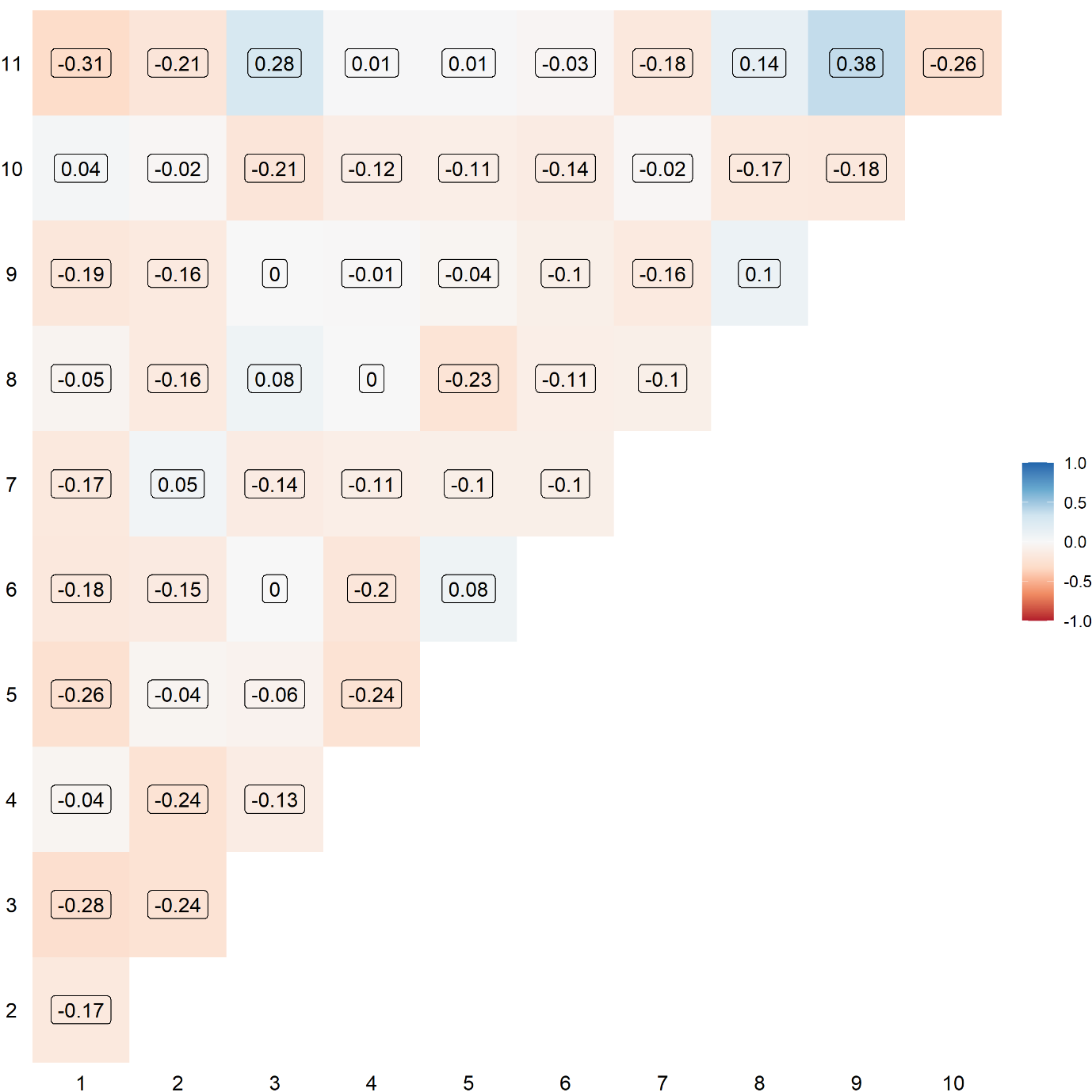

Figure S3: Correlation between topics.

### Tables

Table S1: Government-related keywords and counts. Includes spelling mistakes.

Keywords

government (4902), government's (405), trace (396), govt (372), political (344), governments (300), track (290), politicians (248), cummings (183), scientists (144), scientific (143), politics (142), power (142), boris (128), westminster (123), dominic (103), gov (99), johnson (93), tax (86), authorities (83), taxes (83), tory (80), election (75), authority (63), austerity (62), ministers (50), democracy (49), advised (48), pm (48), minister (43), politically (42), sage (40), conservative (38), tracing (37), advisors (34), labour (33), powers (33), goverment (30), elected (29), cabinet (28), vote (28), voted (26), governmental (25), advise (24), parliament (24), elections (23), governed (23), gov't (19), scientist (19), dido (17), govt's (16), governance (15), elective (14), populist (14), democratic (13), politicised (13), advisers (12), conservatives (12), govern (12), populism (12), sturgeon (12), johnson's (11), taxpayers (11), governement (10), gvt (10), whitty (10), advising (9), authoritarian (9), authoritarianism (9), establishments (9), govenment (9), hancock (9), hmg (9), politician (9), tracked (9), parliamentary (8), pm's (8), politicising (8), taxation (8), voting (8), barnard (7), cummins (7), durham (7), establishment (7), ferguson (7), minister's (7), chancellor (6), goverments (6), politicisation (6), socialist (6), taxpayer (6), totalitarian (6), tracks (6), unelected (6), whitehall (6), adviser (5), chris (5), elect (5), electing (5), governing (5), libertarian (5), thatcher (5), advisor (4), advisory (4), autocracy (4), bojo (4), boris's (4), electorate (4), gove (4), governor (4), govts (4), hmrc (4), phe (4), referendum (4), scientifically (4), taxed (4), tracking (4), votes (4), borris (3), farage (3), governors (3), govnt (3), jenrick (3), politicise (3), politicking (3), populists (3), reelected (3), rightwing (3), rishi (3), sadiq (3), taxing (3), traced (3), voter (3), advises (2), antidemocratic (2), authoritative (2), bureaucracy (2), bureaucratic (2), bureaucrats (2), cabinets (2), democracies (2), democratically (2), electoral (2), givernment (2), gov.com (2), gov.uk (2), goverment's (2), governemtn (2), govmt (2), govn (2), gvmt (2), johnsons (2), johnston (2), libertarians (2), polical (2), politic (2), politicans (2), politicization (2), politicizing (2), socialism (2), sunak (2), sunak's (2), taxiing (2), toryscum (2), tracer (2), tracker (2), undemocratic (2), unscientific (2), westminister (2), antiestablishment (1), antiestablishmentarianists (1), atuhorities (1), auth9orities (1), authoritarians (1), authouritarian (1), autocratic (1), bj (1), burnham (1), burueacracy (1), cabinet's (1), chancellors (1), churchill (1), churchill's (1), conservaties (1), corbyn (1), cummingsgate (1), dcm (1), dcms (1), domini (1), dominick (1), domminic (1), domnic (1), doris's (1), e.g.bojo (1), etonians (1), farage's (1), from.government (1), givenment (1), gobernment (1), goernment (1), gorenment (1), gouvernement (1), govenmental (1), govenments (1), govenrnment (1), governmant (1), governmenet's (1), governmenst (1), government.think (1), governmentresponses (1), governments.this (1), governmentt (1), governmet (1), governmrnt's (1), goverrnment (1), govertment (1), govt.messages (1), gvnt (1), gvts (1), jenryk (1), jonson's (1), lefites (1), leftie (1), liarjohnson (1), parliaments (1), pms (1), policitians (1), politians (1), politicallu (1), politicians.politicians (1), politicitians (1), politicos (1), polititians (1), popularism (1), populistm (1), primeminister (1), priti (1), publichealth (1), reelection (1), rishi's (1), rulers (1), rulership (1), rushi (1), sage's (1), starmer (1), sunack (1), taxpayer's (1), thatcher's (1), thatcherite (1), torygraph (1), totalitarianism (1), tracers (1), westminster's (1), westminsters (1), williamson (1)

Table S2: Number of responses using government-related keyword by question

| Question | Responses |
| --- | --- |
| Q1. Is there anything you would like to tell us about the changes that have been brought about by the Covid-19 pandemic and the impact these have had on your mental health or wellbeing? | 702 |
| Q2. What is bothering you the most about the pandemic? What aspects of it have you been finding most difficult? | 2,701 |
| Q3. Has the pandemic had any negative impacts on your mental health and wellbeing? If so could you tell us about these? | 384 |
| Q4. Has the pandemic had any positive impacts on your mental health and wellbeing? If so could you tell us about these? | 69 |
| Q5. Since the Covid-19 pandemic began, how have you been feeling about the future? What are you hopeful or concerned about? | 1,415 |
